# A cross-country analysis of episodic memory and (potentially) modifiable risk factors of dementia

**DOI:** 10.1101/2024.02.09.24302563

**Authors:** David Knapp, Arie Kapteyn, Alessandro Giambrone, Tabasa Ozawa

## Abstract

The widely cited Lancet Commission concluded that 40% of dementia cases may be preventable through interventions targeting what they refer to as modifiable risk factors. These risk factors have been widely studied individually, but rarely investigated collectively and across many countries. If these factors are “true” (i.e., impactful) modifiable risk factors, then their independent relationship should be robust across countries and comorbidities. We analyze the cross-country consistency of relationships between these modifiable risk factors and episodic memory, a common predictor of cognition and dementia. Using internationally comparable aging studies in 31 countries including the United States, England and Europe, we estimate regressions of combined immediate and delayed word recall with modifiable risk factors and demographic characteristics. Cross-country differences in culture, policies, economy, and other collective experiences lead to significant variation in lifecycle outcomes, including cognitive decline and modifiable risk factors. Our approach does not conclusively affirm a causal relationship but can identify relationships that are weak or nonexistent. We find a limited number of robust relations: education, depression, and hearing loss show clear, consistent associations with our cognition measure. The evidence for other factors, including obesity, smoking, diabetes, and hypertension is weaker and becomes almost non-existent when correcting for multiple hypotheses testing. The inconsistent relationships across countries between episodic memory and obesity, smoking, diabetes, and hypertension suggest the lack of a causal mechanism leading to cognitive decline – a necessary condition for these risk factors to be modifiable and effective targets for policy interventions aimed at controlling dementia prevalence and cost.

## Introduction

The prevalence and cost of dementia are substantial: 55 million people worldwide estimated to rise to 78 million in 2030 and 139 million by 2050 [1]; the global cost of dementia was $263 billion in 2019, and is projected to reach over $1.5 trillion in 2050 [2]. Reducing this upward trend in prevalence and cost is an international public health priority.

The widely cited Lancet Commission concluded that 40% of dementia cases may be preventable, through appropriate interventions targeting what they refer to as modifiable risk factors. These factors include:

1. Education
2. Hearing loss
3. Diabetes
4. Hypertension
5. Obesity
6. Smoking
7. Depression
8. Physical inactivity
9. Social isolation
10. Traumatic brain injury (TBI)
11. Alcohol
12. Air pollution

These factors have been widely studied individually, but rarely investigated collectively and across many countries.

Abating the rise in dementia prevalence and cost requires identifying risk factors that contribute to cognitive decline (i.e., identifying a causal mechanism) and that are modifiable by a person’s behaviors and choices. If these factors are “true” (i.e., impactful) modifiable risk factors, that is they cause differences in cognitive levels, then there should exist a measurable relationship between the risk factor and measures of cognition. Further, the causal relationship should be robust enough that it is measurable when considering other demographic and risk factors and replicable across environmental contexts, such as across different countries.

Our analysis addresses the cross-country consistency of relationships between these modifiable risk factors and cognition. We do not aim to prove these are causal relationships. Rather, we approach the question from the opposite direction – if these relationships are causal, then they should be robust to confounders and be identifiable in many countries. Cross-country differences in culture, policies, economy, and other collective experiences lead to significant variations in lifecycle outcomes, including dementia onset and modifiable risk factors. As we demonstrate, many of these modifiable risk factors are inconsistently associated with our measure of cognition, particularly after accounting for concurrent factors. For factors with inconsistent relationships, this raises the possibility that these factors are more signals of other conditions or dementia itself rather than causal modifiable risk factors of dementia.

We examine the relationship between these modifiable risk factors and a measure of cognitive function by analyzing data from an internationally comparable set of aging studies with nationally representative samples covering the United States (Health and Retirement Study, HRS), most of Europe (Survey of Health, Ageing, and Retirement in Europe, SHARE), and England (English Longitudinal Study of Ageing, ELSA) [3–5]. These studies have been widely used for aging research. We examine the first seven measures – the remaining five (physical inactivity, social isolation, TBI, alcohol, and air pollution) are left out of our analysis due to the limited availability of common measures for these risk factors across surveys at this time. We measure cognitive function using 10-word immediate and delayed word recall, measures of episodic memory, that are consistently asked across surveys. Memory is a key symptom of dementia, and longer word recall tests have a successful history of predicting dementia and cognitive impairment [6,7].

### Past research on modifiable risk factors

We briefly review past research on each risk factor’s likely mechanisms for affecting cognition and potential analytical challenges when assessing the validity of those mechanisms.

#### Education

Higher educational attainment is associated with a lower risk of dementia [8–11]. The cognitive reserve hypothesis suggests that those with higher educational attainment may be more able to delay age-related neurodegeneration, but more research is needed to reveal the precise mechanisms [12]. Mediation through socioeconomic status accounts for some of this association [13]. Additionally, performance on cognitive tests improves with education, raising the possibility of misclassification bias [14].

#### Hearing Loss

Age-related hearing loss in both the peripheral and central auditory systems (detecting and processing sound, respectively) have been associated with an increased risk of dementia [15]. Two common, non-mutually exclusive theories on the mechanism for this relationship exist. The sensory deprivation hypothesis proposes that reduced sensory input leads to social isolation and structural changes in the brain that are associated with dementia [16]. The cognitive load hypothesis posits that the increased cognitive resources required for auditory processing diminish the resources required for cognitive processing [17]. Correlations with dementia may arise from unmeasured confounders such as vascular disease and inflammation as well as testing bias due to difficulties with hearing instructions. More research is needed on the impact of hearing aid use on dementia risk, but a recent randomized control trial comparing hearing and health education interventions found that hearing treatment reduced global cognitive decline among participants with greater cognitive impairment risk at baseline but not among healthy participants [18].

#### Diabetes

Both type 1 and type 2 diabetes increase the risk of dementia, and a younger age at the onset of diabetes is associated with a greater risk [19]. The link between diabetes and dementia is complex, but possible pathways include the increased risk of vascular disease, insulin resistance, hyperglycemia, impaired amyloid beta clearance, hyperphosphorylation of tau protein, oxidative stress, and inflammation [20]. A healthy diet and physical exercise are recommended for diabetes management and have been associated with slowing cognitive decline [21,22]. There is limited evidence on the efficacy of antidiabetic medications and intranasal insulin administration in lowering the risk of dementia [23,24].

#### Hypertension

There is strong evidence that high blood pressure leads to vascular dementia via cerebrovascular disease [25]. Hypertension exhibits an age-dependent association with dementia, with the greatest increase in risk observed in those with midlife hypertension, as well as those who have persistent hypertension from mid to late life [26]. Late-onset hypertension seems to have no significant association with dementia and may even have a protective effect for those over the age of 80 [27]. Analysis of the impact of antihypertensive medication use remains inconclusive [28].

#### Obesity

Mid-life obesity is associated with higher dementia risk, potentially through the development of cerebrovascular disease, an increase in inflammation, and changes in the brain structure that elevate dementia risk [29]. In contrast, late-life obesity seems to exert a protective effect on dementia, an association that may be influenced by survival bias, although other mechanisms have been proposed, such as cognitive enhancement by the hormone leptin which is secreted by fat tissues and overexpressed in genes of obese individuals [30,31]. Being underweight in late life has also been associated with a higher risk of dementia, potentially signaling, rather than causing, dementia [32].

#### Smoking

Current smoking has been associated with an increased risk of dementia but not former smoking [33,34]. Plausible mechanisms include oxidative stress and inflammation that increases amyloid beta concentrations, and increased risk of vascular disease which leads to vascular dementia [35]. In studying the smoking-dementia association, it is necessary to account for the competing risk of death and the selection of “healthy smokers” who survive to old age and may have unmeasured factors that could be protecting them from cognitive decline [9,36].

#### Depression

Current and recent depression have been associated with a higher likelihood of incident dementia compared to no depression and remitted depression [37]. Depression may increase the risk of vascular dementia through its association with chronic inflammation, high blood pressure, small vessel disease, white matter hyperintensities, and reduced blood flow in the brain [38]. Reverse causation could arise if a dementia diagnosis or circumstances around cognitive decline trigger depressive symptoms [39]. There is limited evidence on the effectiveness of antidepressants in lowering the risk of dementia [40].

#### Implications

The research discussed above documents several physiological connections that may be plausible mechanisms for how the identified modifiable risk factors effect cognition. It also reveals that many of the relationships, as measured, have empirical limitations that raise questions about whether there exists a causal mechanism leading to cognitive decline. Since an experimental design across countries is infeasible, we examine the consistency of relationships between the potentially modifiable risk factors and episodic memory (a key proxy for cognition) across countries, reflecting a wide variety of settings. If these factors are “true” (i.e., impactful) modifiable risk factors, then their independent relationship should be robust across countries and comorbidities. Our approach does not conclusively affirm a causal relationship but can identify relationships that are likely weak or nonexistent.

## Methods

### Data

We use data from nationally representative samples of the population age 50 and older from HRS, SHARE, and ELSA for a total of 31 countries in our analysis. All three studies are longitudinal, interview the same respondents biannually, and collect substantial information on respondents and their spouses, including health, cognition, and behaviors. We use data from a respondent’s first interview: there is one observation per respondent, and observations are derived from different interview waves, however the questions used are largely the same. A key motivation for using the respondent’s first interview is that research has found that questions aimed at eliciting cognitive function exhibit improved test scores with re-testing [41]. Sample sizes range from 529 in Cyprus to 35,409 in the US, with a total of 173,020 first interviews across the 31 countries. Supplementary Appendices A and B provide additional detail on our data, including country and sample selection, summary statistics, and regression results that are relevant for readers interested in the studies, specific questions asked, or wanting to replicate our analyses.

### Measures

Our measure of cognitive function is the sum of words recalled from an immediate test and a delayed test in each survey. In these tests, an interviewer reads a list of 10 words, and the respondent lists as many words as they can remember (immediate recall). This exercise is repeated after a few questions on a different topic, taking about 5 minutes (delayed recall). The word lists vary by study, but are common, simple words. Each respondent’s scores are added to create a summary score, which we refer to as the total word recall (TR) score.

Measures of modifiable risk factors are self-reported and include years of education, censored at 17 years; hearing ability (possibly when using a hearing aid) on a five-point scale; whether the respondent has been told by a doctor that they have: diabetes or high blood sugar, or high blood pressure or hypertension; a constructed categorical measure of being either obese (body mass index at or above 30), underweight (less than 18.5), or neither based on reported height and weight; a constructed categorical measure of smoking based on whether they report currently being or ever having been a smoker, defined by a smoking period of at least one year; and for depression, a mental health index ranging from 0 to 12 in SHARE, based on EURO-D, and 0 to 8 in HRS and ELSA, based on CESD (see Appendix A.2 for details on differences in survey questions across time and study), that consider a respondent’s recent feelings of depression, sadness, happiness, and loneliness, among other indicators of mental health.

Risk factors are measured on different scales making comparisons of effect sizes difficult. To facilitate interpretation, we standardize each continuous risk factor (except binary and categorical variables), by country, so that the coefficient estimates are expressed in terms of their standard deviation or as the effect of the presence of a risk factor within each country (i.e., we compute z-scores). Additionally, we standardize TR scores to make the interpretation of the results across countries comparable since interview waves 1 and 2 of HRS provided 20 words instead of 10. We standardize within each interview wave the immediate and delayed recall score, and then sum them together to obtain the TR score, as a way to account for this difference in how the variable is measured. The score is then normalized once again across all waves. This process is discussed in detail in Appendix A.3.

We maintain more than 97% of the original sample that has a TR score at their first interview in each country (Appendix A.4 discusses sample restrictions). Appendix A.5 summarizes sample means for the TR score, risk factors, and remedies. A key takeaway from these summary statistics is that there exist significant cross-country differences in risk factor experience and remedy use in the 31 countries included in our analysis. For example, the average duration of respondents’ education ranges from 6.3 years in Portugal to 12.9 years in Denmark.

### Regression models

For each modifiable risk factor and country, we estimate a simple bivariate model (Model 1) where we regress TR on each risk factor separately and a complete model (Model 2) where we include all risk factors as covariates in the regression model, plus a variable for sex, and a categorical variable for age (age groups: 50-59, 60-69, 70-79, 80+). Model 2’s specification is:

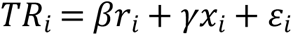

where *TR_i_* is the standardized total word recall score for respondent *i* the first time he or she was interviewed; *r_i_* is the vector of standardized or binary risk factors and *x_i_* is a vector of background variables; *ε_i_* is the error term. We estimate each model separately by country.

We consider one extension to Model 2. We investigate the role of a risk factor’s possible remedy (e.g., taking blood pressure medication, quitting smoking) in abating the relationships between TR and the risk factor.

When presenting results, we report model coefficients from ordinary least squares regression on the standardized TR measure using robust standard errors. Our analysis was conducted using STATA 16.1.

## Results

### Relationship consistency between risk factors and cognitive function

Models 1 and 2 are estimated separately by country to allow the estimated relationships to differ in magnitude (i.e., the relationships do not have to be identical across countries). Figure 1 presents the cross-country estimates for years of education (panel A), high blood pressure (panel B), and current smoking (panel C). The vertical axis is the coefficient of years of education on cognition (as measured by total word recall). The dotted line is the cross-country average.

**Figure 1:**
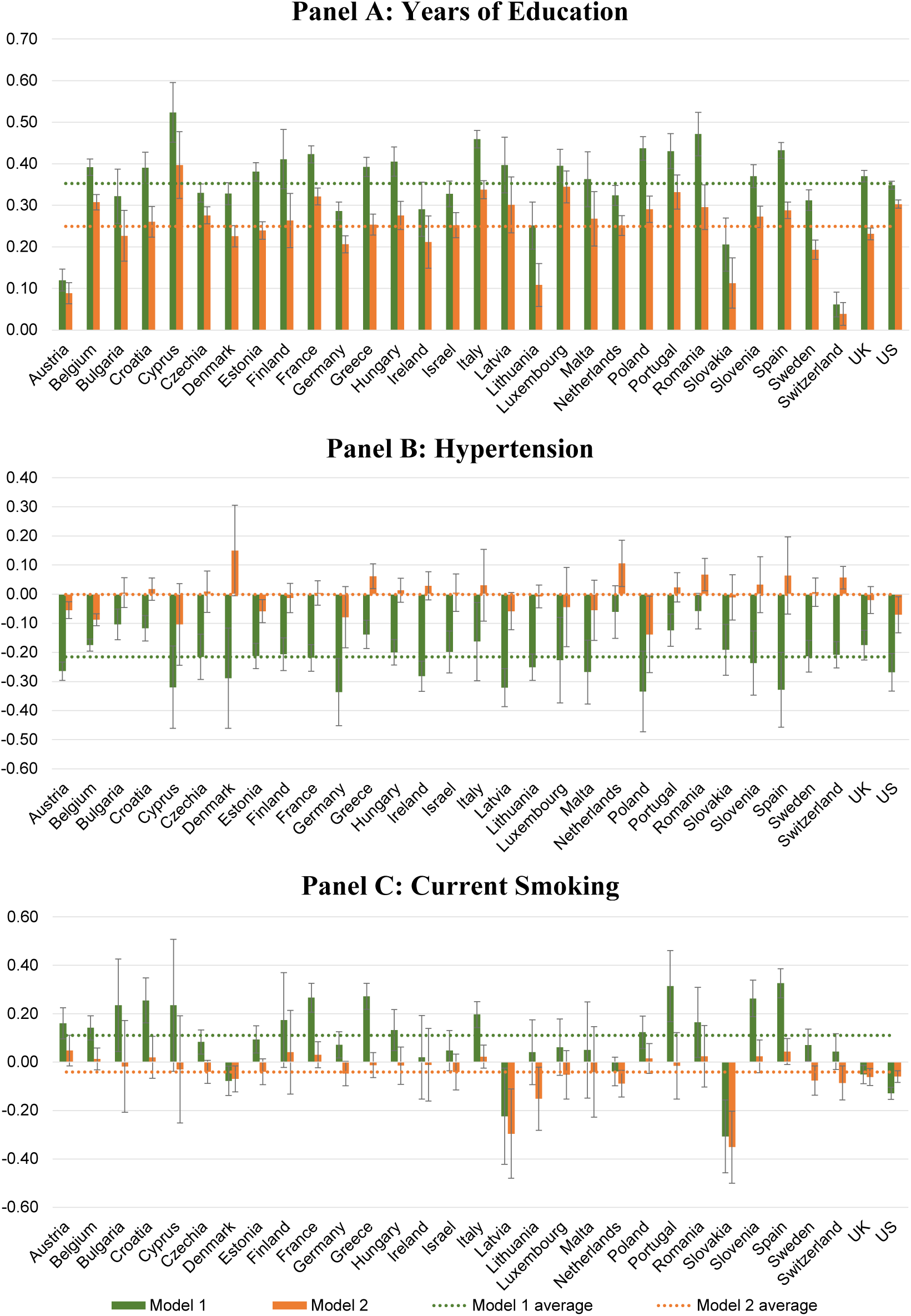
Standardized coefficients by country from regressing TR on risk factors

We find a strong positive relationship between education and cognition. An increase of one standard deviation (sd) in years of education was associated with an average 0.35 sd increase across the 31 countries in TR. This relationship persists accounting for other risk factors, age, and sex. The average relationship across the 31 countries is 0.25 sd in Model 2. For some countries the relationship is notably smaller, but in all countries it is significantly different from zero. This indicates a consistently significant relationship between education and cognition.

In Figure 1, Panel B, we find a consistent negative relationship for Model 1, which shows the relationship between hypertension and cognition not accounting for comorbidities. The relationship is significant and negative for 29 of the 31 countries (94%) with a cross country average of −0.22 sd. However, accounting for age, sex, and other risk factors in model 2, we find no relationship on average, with only 5 countries having significant negative relationships (Austria, Belgium, Estonia, Poland, US), and 4 having significantly positive relationships (Greece, Netherlands, Romania, Switzerland). This suggests a relationship that is not consistent with this risk factor determining cognitive levels at older ages.

In Figure 1, Panel C, we counterintuitively find many countries with positive relationships for Model 1, which suggests current smoking is associated with higher cognition levels. The relationship is significant and positive for 17 of the 31 countries (55%) with a cross country average of +0.11 sd. Only four countries have the expected significant, negative relationship. Accounting for age, sex, and other risk factors in Model 2, we find a slight negative relationship on average (−0.04 sd), with 9 (29%) countries having significant negative relationships. This too suggests a relationship that is not consistent with this risk factor determining cognitive levels at older ages.

Figure 2 summarizes the standardized coefficients for the risk factors, arranged from largest to smallest relationship with cognitive function based on the simple model (Model 1). Figure 2 presents the US as a separate counterpoint to the cross-country average as many existing studies use a US-based sample.

**Figure 2:**
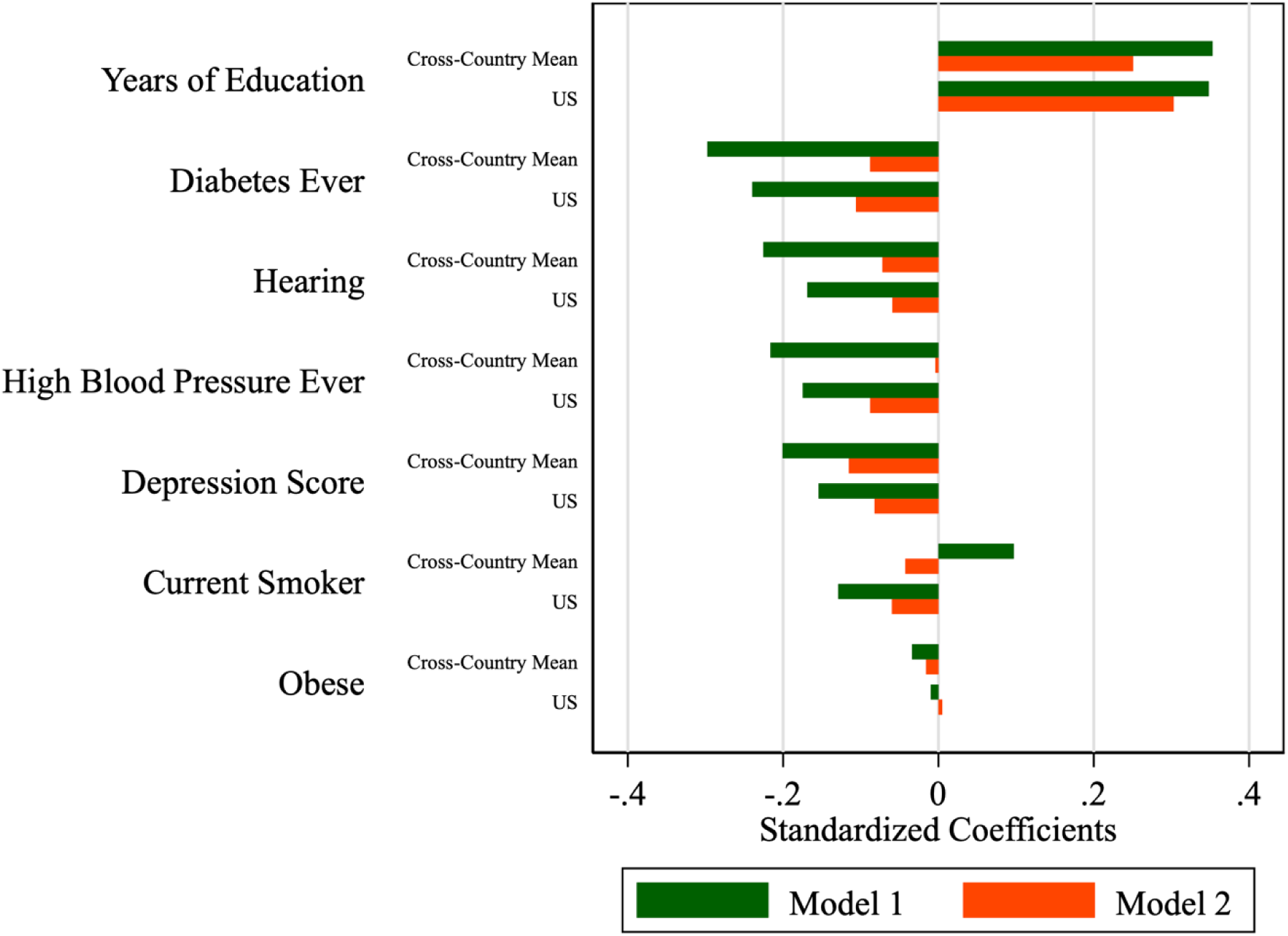
Standardized coefficients from regressing TR on risk factors, cross-country mean and US value. Notes: Risk factors are sorted by the largest to smallest based on the cross-country mean of the absolute standardized coefficient in Model 1.

Table 1 summarizes the proportion of countries that have a positive or negative sign and how many coefficients are statistically significant. In Table 1, we also report how many coefficients are statistically significant following a Bonferroni correction for multiple hypotheses testing. With 31 countries, there is an 80% (1 ― 0.95^31^ = 0.80) probability of at least one Type 1 error (i.e., false positive), where statistical significance is judged at the 5% level. Our conservative correction is to require statistical significance at the 5/31 = 0.161% level in each country. In this way, the joint probability of a Type 1 error is only 5% (1 ― (1 ― 0.00161)^31^ = 0.05.

**Table 1:**
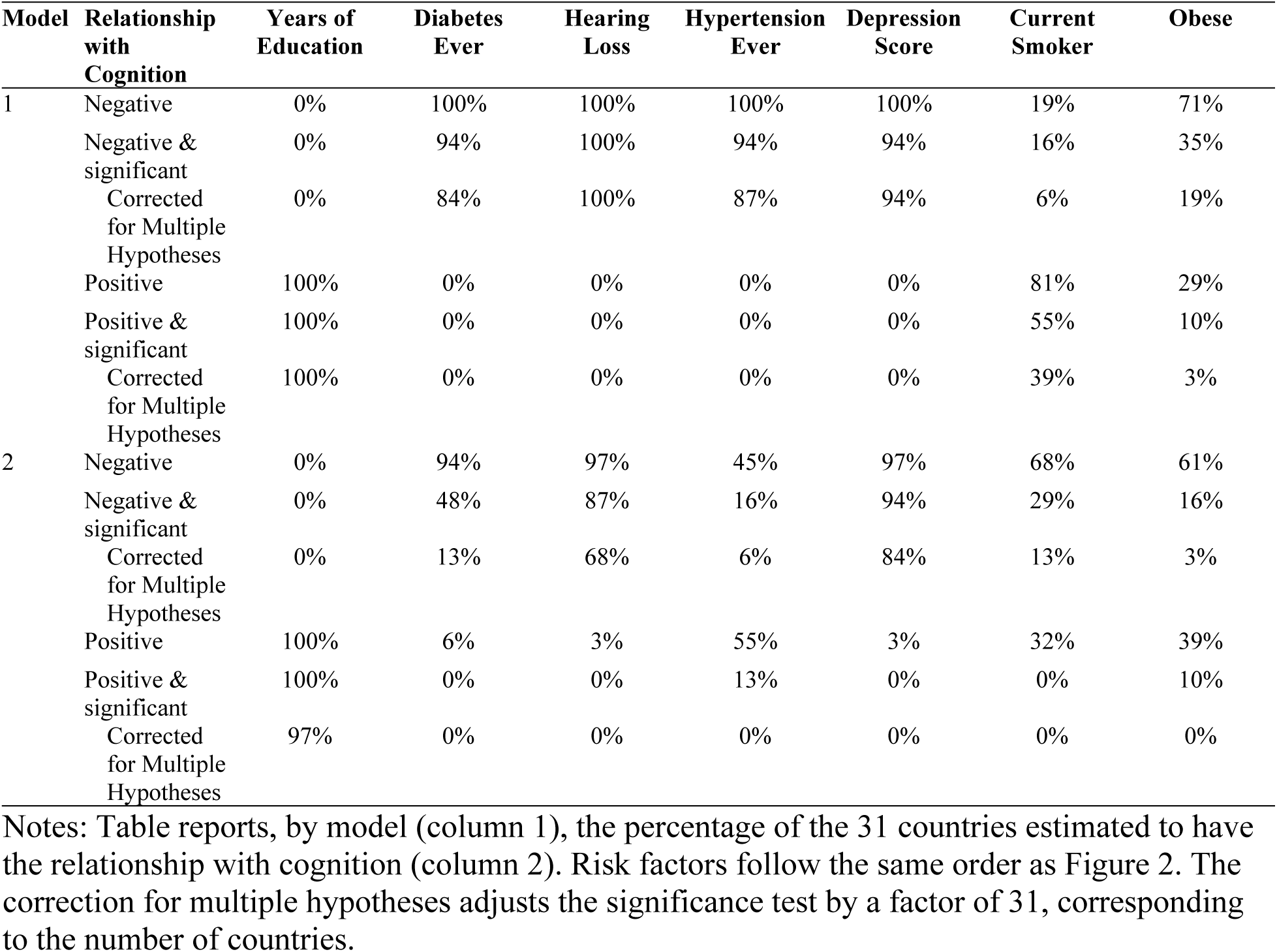
Distribution of coefficients across countries between positive, negative, positive and significant, and negative and significant.

We define a risk factor as having a consistent cross-country relationship with cognitive levels (as measured by TR score) if at least half of the countries have significant common directional relationships and no countries having significant reverse relationships. The bivariate relationships between the risk factors and cognitive function are consistent with relationships in Livingston et al.’s (2017) life-course model with education, hearing loss, and depression having substantial and significant relationships. As noted above, an increase of one sd in years of education was associated with an average 0.35 sd increase across countries in TR (USA: 0.35 sd), with all relationships being positive and statistically significant. An increase of one sd in difficulty hearing or the depression score was associated with an average 0.23 and 0.20 sd decrease in TR (USA: 0.17 and 0.15 sd), respectively, with all relationships being negative and statistically significant.

Using the simple model, we also find a substantial relationship across countries for the comparatively less impactful risk factors in Livingston et al. (2017). Ever having diabetes, hypertension, or being obese were associated with an average 0.30, 0.22, and 0.03 sd decrease in TR (USA: 0.24, 0.17, 0.01 sd), respectively, and a substantial majority (74% or more) of countries show negative and statistically significant relationships. In contrast with what one would expect based on Livingston et al.’s (2017) model, current smoking was associated with an average *increase* in TR (0.10 sd) and was positive for 81% of countries in our analysis and statistically significant for 55% of the countries. For the USA, the simple model is associated with a statistically significant 0.13 sd decrease in TR, which is consistent with Livingston et al. (2017) and more recent research suggesting smoking has a negative impact on cognitive function.

Accounting for sex, age, and the comorbid risk factors (Model 2), the relationship between cognitive function and risk factors substantially diminishes in magnitude. Among the risk factors with substantial and significant relationships in Livingston et al. (2017) – education, hearing loss, and depression – we found the relationships remain consistent in direction and statistically significant for a substantial majority of countries, but the magnitude of the relationship is smaller. For years of education, a one sd in years of education was associated with an average 0.25 sd increase across countries in TR, about 30% smaller than in the simple model. An increase of one sd in difficulty hearing or the depression score was associated with an average 0.07 and 0.12 sd decrease in TR, about 70% and 40% smaller than the magnitude in the simple model, respectively. For years of education, hearing loss, and depression, the findings are statistically significant in 97%, 68%, and 84% of countries even with our correction for multiple hypotheses.

Accounting for comorbidities markedly reduces the negative relationships across countries for ever having diabetes, hypertension, or being obese, and those relationships are only negative and statistically significant for 48%, 16%, and 16% of the countries, respectively. Correcting for multiple hypotheses, we find that the relationship between TR score and these risk factors is robust for 13% or fewer countries. A minority of countries (13%) exhibit positive and statistically significant relationships between ever having had hypertension and TR, but none of these relationships are robust to our correction for multiple hypotheses.

For those currently smoking, 68% of countries exhibit a negative coefficient, compared to only 19% in the simpler model. However, only 29% of countries exhibit negative and significant relationships, and this shrinks to 13% after correcting the significance levels for multiple hypotheses. For these risk factors, the relationship with TR may not be fully captured by our model, or the relationship is weak or non-existent.

Summarizing, we find inconsistency across countries in the relationship between cognitive function and hypertension, diabetes, obesity, and smoking. These findings suggest that these relationships may be reflecting omitted factors that vary by country, rather than representing a risk factor where modifications to behavior may potentially alter the trajectory of cognitive aging. In Online Appendix C, we consider a couple of supplementary analyses:

1. Does accounting for the age of diabetes diagnosis affect our finding that diabetes is inconsistently associated with our cognition measure?
2. Does the relationship with risk factors change with age?

We address the first question by accounting for differential timing of diabetes diagnosis, and the second question by allowing for the relationship between risk factors and our measure of cognition to vary by age. Neither supplementary analysis substantively changes our findings.

### Do risk factor remedies reduce negative relationships with cognitive function?

We consider an extension of Model 2 that incorporates remedies for potential risk factors as additional covariates. Figure 3 compares the coefficients from the risk factors to the coefficients on the remedies themselves (coefficients in Figure 3 may differ from Figure 2 because of the extended model).

**Figure 3:**
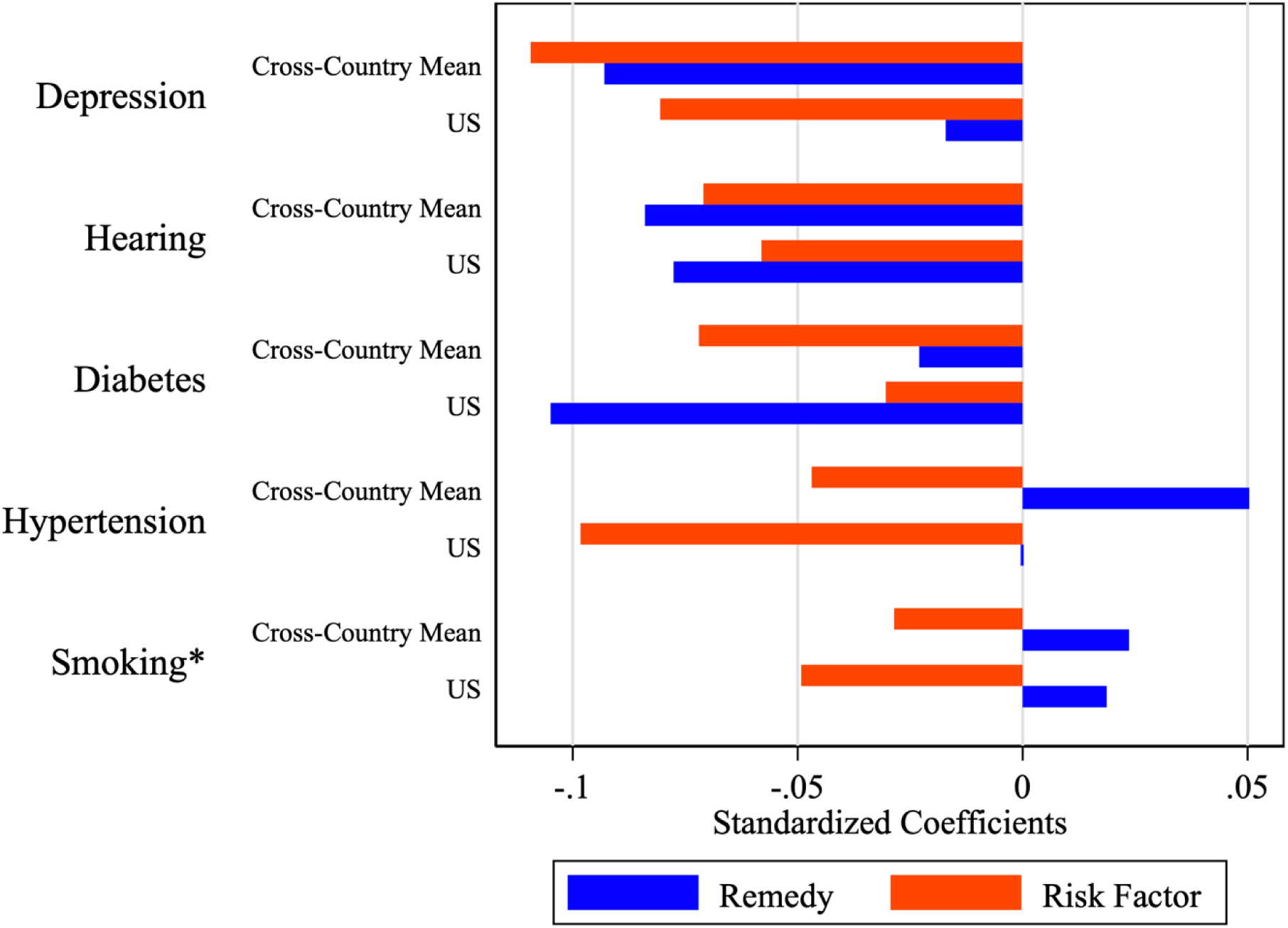
Standardized coefficients of the risk factors and their respective remedies, cross-country mean and US value. Notes: Reported coefficients are based on an extension to Model 2 that adds indicator variables for whether a respondent takes medication for diabetes, depression, or hypertension, has a hearing aid, or is a former smoker. In most cases, the coefficients are added to determine the net effect on the TR score, that is the blue bars are the sum of the effect of having a condition and the separate effect of using a remedy. The exception is smoking (*), where the remedy (being a former smoker) is mutually exclusive to the risk factor (i.e., current smoker). In this case, the net effect on the TR score of the remedy is just the remedy’s standardized coefficient.

With the exception of smoking and quitting (smoking’s remedy), the coefficients are additive: if a respondent has the risk factor and uses the remedy, then the total effect is the sum of the red and blue bars. If a remedy reduces the relationship between the risk factor and cognitive decline, then the blue bar will point in the opposite direction of the red bar. However, this is only the case for the country mean of taking medications for hypertension. In all other cases, the blue and red bars point in the same direction (we discuss the case of smoking separately below). In the case of hypertension, the country mean suggests that having the risk factor and using the remedy (i.e., having high blood pressure and taking medication) is as good as not having the condition (the blue and red bars point in opposite directions, and are of equal length). However, that does not appear to be the case for the US, where the remedy has minimal effect.

For diabetes, hearing loss, and depression the figure suggests that the remedy makes the effect on memory worse. A plausible interpretation is that the use of a remedy is an indicator of the severity of a condition. Hearing loss is a case in point. The question asks if a respondent has difficulty hearing, also when wearing a hearing aid. Having difficulty hearing with a hearing aid likely implies worse hearing than having difficulty without a hearing aid. The interpretation for diabetes and depression is less straightforward, but it still seems plausible that medication is more likely to be prescribed if the condition is more severe.

As noted, the bars in Figure 3 should be interpreted differently for smoking. The reference category is never having smoked. The red bars indicate memory for current smokers, while the blue bars indicate former smokers. Thus it appears that former smokers actually score better than never smokers. This pattern hides considerable heterogeneity across countries. Current smoking has a significant negative relationship with the TR score in the US, but this relationship is not significantly different from a null effect in other countries, with the exception of England, Latvia, the Netherlands, and Slovakia (for Austria, France, and Spain, it is significantly positive). If past smoking was associated with lower cognition at older ages, we would have expected to observe a negative coefficient for the remedy. However, only in Romania is there a statistically significant negative relationship between past smoking and TR score. A significant positive relationship exists in 10 of 31 countries (including Austria, Belgium, France, Italy, and Spain).

Three possible interpretations of this relationship are that the act of quitting smoking could either lead to greater cognitive levels in old age (a causal interpretation), smoking cessation could lead to the adoption of other healthy behaviors that are beneficial for cognition such as exercise and healthy eating (a mediation interpretation), or the estimated relationships could indicate that people with better cognition are more likely to quit (a selection interpretation).

Overall, while the majority of modifiable risk factors have known remedies, with the exception of hypertension and smoking, we do not find evidence that the remedies reduce the negative relationship between these risk factors and measured cognition. In the case of hypertension and smoking, as noted in Table 1, the relationship between the risk factor and TR score is inconsistent and not typically significantly different from a null effect. For factors that are consistently related to TR score (e.g., depression, hearing loss), our results suggest there is sorting into the use of the remedy based on the severity of the condition, leading to limited insights from point-in-time data. To test whether remedies for those risk factors are causally able to limit cognitive decline, a longitudinal experiment is required.

## Discussion

Establishing causal patterns affecting dementia risk at older ages is challenging based on observational data. On the other hand conducting randomized controlled trials that experimentally vary factors that are thought to influence dementia risk are generally infeasible, except in limited interventions aimed at slowing cognitive decline. Many factors affecting dementia risk are likely to exert their influence over long periods. Examples include education, early life conditions, occupational experiences, and pollution. Establishing causal effects of experimental manipulations of such factors would likely take several decades, and in most cases would raise grave ethical concerns. This leaves us with exploiting natural experiments, such as changes in compulsory schooling laws [43,44]. Another source of arguably exogenous variation can be found in international variations in institutions and policies.

This paper exploits harmonized data from 31 countries. Before addressing the effects of policy and institutional variation, the first step is to investigate the robustness of associations proposed in the literature. Taking the widely cited Lancet Commission report as our starting point, we only find a limited number of robust relations: education, depression, and hearing loss show clear associations with our cognition measure, the sum of immediate and delayed recall. The evidence for the other factors is weaker and becomes almost non-existent when correcting for multiple hypotheses testing.

The lack of robustness may have different reasons. The measures used in the study may not be perfectly comparable across countries, but in view of the harmonized nature of the surveys from the different countries, that is likely to play a relatively minor role. It seems more likely that several of the hypothesized relations at least partly proxy for omitted factors. For example, obesity or smoking reflect lifestyles and personal circumstances that may affect dementia risk beyond these two observed factors. The correlation between the observed and unobserved factors is likely to differ across countries. For example, smoking rates vary across countries, and so does its relation with socio-economic background. If smoking is a proxy for these other background variables, then we expect the observed relation between smoking and memory to vary across countries. And that is precisely what we find.

## Limitations

The primary limitation of this study is undoubtedly that it is an observational study, but as noted, in many ways that is inevitable. The study uses only cross-sectional data, even though the underlying datasets are all panels. The reason for only using data from the first time a respondent answers a survey is to avoid practice effects. Additionally, because cross-sectional data cannot establish temporality, it is susceptible to reverse causality. For example, it is possible that the significant and negative association between depression and memory may have been a result of depression being a prodrome, or early symptom, of dementia. Our analysis is not intended to conclusively affirm a causal relationship but identifies relationships that are weak or nonexistent. Finally, we do not have a measure of dementia diagnosis, rather use measures of memory as a proxy for cognitive function. Clinical assessments of dementia are not practical in large panel surveys, like those used in our analysis. Risk factors may contribute to cognitive decline through alternative pathways other than memory (e.g., attention or executive functions, such as reasoning).

## Data Availability

All data used in the study is public use without cost and may be downloaded by registering at the respective study's website:
Health and Retirement Study: https://hrs.isr.umich.edu/data-products -
English Longitudinal Study of Ageing: https://beta.ukdataservice.ac.uk/datacatalogue/series/series?id=200011
Survey of Health, Ageing, and Retirement in Europe: https://share-eric.eu/data/become-a-user

https://beta.ukdataservice.ac.uk/datacatalogue/series/series?id=200011

https://share-eric.eu/data/become-a-user

## Acknowledgments

We thank the study participants and coordinators of ELSA, HRS, and SHARE for providing valuable data for our study.

## Supporting information

S1 Appendix

